# Weighted burden analysis of rare coding variants in 470,000 exome-sequenced UK Biobank subject characterises effects on hyperlipidaemia risk

**DOI:** 10.1101/2023.10.19.23297272

**Authors:** David Curtis

## Abstract

A previous study of 200,000 exome-sequenced UK Biobank participants investigating the association between rare coding variants and hyperlipidaemia had implicated four genes, *LDLR*, *PCSK9*, *APOC3* and *IFITM5*, at exome-wide significance. In addition, a further 43 protein-coding genes were significant with an uncorrected p value of <0.001. Exome sequence data has become available for a further 270,000 participants and weighted burden analysis to test for association with hyperlipidaemia was carried out in this sample for the 47 genes highlighted by the previous study. There was no evidence to implicate *IFITM5* but *LDLR*, *PCSK9*, *APOC3*, *ANGPLT3*, *ABCG5* and *NPC1L1* were all statistically significant after correction for multiple testing. These six genes were also all exome-wide significant in the combined sample of 470,000 participants. Variants impairing function of *LDLR* and *ABCG5* were associated with increased risk whereas variants in the other genes were protective. Variant categories associated with large effect sizes are cumulatively very rare and the main benefit of this kind of study seems to be to throw light on the molecular mechanisms impacting hyperlipidaemia risk, hopefully supporting attempts to develop improved therapies.

This research has been conducted using the UK Biobank Resource.

## Introduction

A previous weighted burden analysis of rare coding variants observed in 200,000 exome-sequenced UK Biobank participants implicated four protein-coding genes as being involved in risk of hyperlipidaemia at exome-wide significance: *LDLR*, *PCSK9*, *ANGPTL3* and *IFITM5* (Curtis, 2021). For all of these except *LDLR*, rare variants predicted to impair gene functioning were associated with lower risk of hyperlipidaemia. It was noted that 55 genes, of which 47 were protein-coding, had uncorrected p values significant at p < 0.001 whereas only 23 would be expected by chance and a number of these genes seemed to be plausible biological candidates. Exome sequence data for a full set of 470,000 participants has now been released and a follow-up study was carried out in order to determine if any of these 47 genes of interest demonstrated evidence for association in the 270,000 newly available samples. For such genes, further analyses were carried out in the whole sample to investigate the overall evidence for association and the contributions from different types of variant.

Most of this exome sequence data has also been used in two previous studies which tested for gene-wise associations with very large numbers of phenotypes, including some hyperlipidaemia-related phenotypes (Backman *et al*., 2021; Wang *et al*., 2021). It was recognised that results from the current investigation would need to be considered in this context of these studies.

## Methods

The same methods were used largely as described in previous studies and the description is repeated here for convenience.

UK Biobank participants are volunteers intended to be broadly representative of the UK population and are not selected on the basis of having any health condition. UK Biobank had obtained ethics approval from the North West Multi-centre Research Ethics Committee which covers the UK (approval number: 11/NW/0382) and had obtained informed consent from all participants. The UK Biobank approved an application for use of the data (ID 51119) and ethics approval for the analyses was obtained from the UCL Research Ethics Committee (11527/001). The UK Biobank Research Analysis Platform was used to access the Final Release Population level exome OQFE variants in PLINK format for 469,818 exomes which had been produced at the Regeneron Genetics Center using the protocols described here: https://dnanexus.gitbook.io/uk-biobank-rap/science-corner/whole-exome-sequencing-oqfe-protocol/protocol-for-processing-ukb-whole-exome-sequencing-data-sets (Backman *et al*., 2021). All variants were then annotated using the standard software packages VEP, PolyPhen and SIFT (Kumar, Henikoff and Ng, 2009; Adzhubei, Jordan and Sunyaev, 2013; McLaren *et al*., 2016). To obtain population principal components reflecting ancestry, version 2.0 of *plink* (https://www.cog-genomics.org/plink/2.0/) was run with the options *--maf 0.1 --pca 20 approx* (Chang *et al*., 2015; Galinsky *et al*., 2016).

The hyperlipidaemia phenotype was determined in the same way as previously from four sources in the dataset: self-reported high cholesterol; reporting taking cholesterol lowering medication; reporting taking a named statin; having an ICD10 diagnosis for hyperlipidaemia in hospital records or as a cause of death (Curtis, 2020a). Participants in any of these categories were deemed to be cases with hyperlipidaemia while all other subjects were taken to be controls. In the primary analyses to implicate specific genes, attention was restricted to participants not included in the earlier study, consisting of 97,050 cases and 207,216 controls. For the subsequent analyses using the whole sample there were 106,091 cases and 363,674 controls.

The SCOREASSOC program was used to carry out a weighted burden analysis to test whether, in each gene, sequence variants which were rarer and/or predicted to have more severe functional effects occurred more commonly in cases than controls (Curtis, 2012, 2016, 2020b). Attention was restricted to rare variants with minor allele frequency (MAF) <= 0.01 in cases or controls or both. As previously described, variants were weighted by overall MAF so that variants with MAF=0.01 were given a weight of 1 while very rare variants with MAF close to zero were given a weight of 10. Variants were also weighted according to their functional annotation using the GENEVARASSOC program, which was used to generate the input files for weighted burden analysis by SCOREASSOC. Variants predicted to cause complete loss of function (LOF) of the gene were assigned a weight of 100. Nonsynonymous variants were assigned a weight of 5 but if PolyPhen annotated them as possibly or probably damaging then 5 or 10 was added to this and if SIFT annotated them as deleterious then 20 was added. The full set of weights and categories is displayed in Table 1 of the previous study (Curtis, 2021). As described previously, the weight due to MAF and the weight due to functional annotation were multiplied together to provide an overall weight for each variant.

**Table 1.**

| **Symbol** | **SLP in original sample** | **Name** | **SLP in new sample** | **SLP in combined sample** |
| --- | --- | --- | --- | --- |
| *LDLR* | 50.08 | Low Density Lipoprotein Receptor | 87.28 | 156.81 |
| *G6PC* | 5.55 | Glucose-6-Phosphatase Catalytic Subunit | 0.67 |  |
| *SULT1E1* | 4.63 | Sulfotransferase Family 1E Member 1 | 0.57 |  |
| *SLC35G1* | 4.38 | Solute Carrier Family 35 Member G1 | -0.06 |  |
| *PLA2G5* | 4.15 | Phospholipase A2 Group V | 1.05 |  |
| *CMTM7* | 3.99 | CKLF Like MARVEL Transmembrane Domain Containing 7 | -0.34 |  |
| *DEFB131A* | 3.66 | Defensin Beta 131A | 0.43 |  |
| *OTULIN* | 3.62 | OTU Deubiquitinase With Linear Linkage Specificity | -1.10 |  |
| *FAM122C* | 3.51 | Family With Sequence Similarity 122C | 0.36 |  |
| *CMIP* | 3.48 | C-Maf Inducing Protein | 0.02 |  |
| *EIF4B* | 3.45 | Eukaryotic Translation Initiation Factor 4B | 0.52 |  |
| *PPP2R3B* | 3.41 | Protein Phosphatase 2 Regulatory Subunit B"Beta | 0.17 |  |
| *HNRNPAB* | 3.38 | Heterogeneous Nuclear Ribonucleoprotein A/B | -0.30 |  |
| *PREB* | 3.37 | Prolactin Regulatory Element Binding | 0.93 |  |
| *PEX12* | 3.36 | Peroxisomal Biogenesis Factor 12 | -0.12 |  |
| *FAM167A* | 3.35 | Family With Sequence Similarity 167 Member A | 0.84 |  |
| *ABCG5* | 3.31 | ATP Binding Cassette Subfamily G Member 5 | 5.08 | 6.95 |
| *ABCD1* | 3.26 | ATP Binding Cassette Subfamily D Member 1 | 0.18 |  |
| *PRAF2* | 3.21 | PRA1 Domain Family Member 2 | -0.45 |  |
| *CTHRC1* | 3.16 | Collagen Triple Helix Repeat Containing 1 | -0.33 |  |
| *SLC25A37* | 3.14 | Solute Carrier Family 25 Member 37 | 0.26 |  |
| *CT62* | 3.11 | Cancer/Testis Associated 62 | 0.41 |  |
| *L1TD1* | 3.09 | LINE1 Type Transposase Domain Containing 1 | 0.02 |  |
| *PIK3R6* | 3.09 | Phosphoinositide-3-Kinase Regulatory Subunit 6 | 1.16 |  |
| *FOXO3B* | 3.08 | Forkhead Box O3B | -0.24 |  |
| *FAM47A* | 3.06 | Family With Sequence Similarity 47 Member A | 0.19 |  |
| *GCK* | 3.04 | Glucokinase | 1.42 |  |
| *MAPKAPK2* | 3.02 | MAPK Activated Protein Kinase 2 | -0.21 |  |
| *PCSK9* | -10.42 | Proprotein Convertase Subtilisin/Kexin Type 9 | -29.08 | -43.57 |
| *IFITM5* | -5.86 | Interferon Induced Transmembrane Protein 5 | 0.44 |  |
| *ANGPTL3* | -5.67 | Angiopoietin Like 3 | -7.48 | -12.68 |
| *APOC3* | -4.89 | Apolipoprotein C3 | -11.01 | -13.19 |
| *PPP1R3G* | -4.25 | Protein Phosphatase 1 Regulatory Subunit 3G | -0.67 |  |
| *TBC1D8* | -3.93 | TBC1 Domain Family Member 8 | -0.90 |  |
| *CTXN2* | -3.91 | Cortexin 2 | -0.64 |  |
| *NPC1L1* | -3.7 | NPC1 Like Intracellular Cholesterol Transporter 1 | -3.42 | -7.60 |
| *ANGPTL4* | -3.66 | Angiopoietin Like 4 | -1.95 | -4.79 |
| *SNX17* | -3.62 | Sorting Nexin 17 | -0.73 |  |
| *SV2B* | -3.29 | Synaptic Vesicle Glycoprotein 2B | 0.23 |  |
| *ITM2B* | -3.23 | Integral Membrane Protein 2B | 0.40 |  |
| *UBR4* | -3.23 | Ubiquitin Protein Ligase E3 Component N-Recognin 4 | 0.82 |  |
| *TXNL4A* | -3.22 | Thioredoxin Like 4A | 0.11 |  |
| *TTR* | -3.16 | Transthyretin | -0.38 |  |
| *GFPT1* | -3.1 | Glutamine--Fructose-6-Phosphate Transaminase 1 | 0.09 |  |
| *APPBP2* | -3.08 | Amyloid Beta Precursor Protein Binding Protein 2 | 0.29 |  |
| *CRYZL1* | -3.06 | Crystallin Zeta Like 1 | -0.70 |  |
| *HLA-A* | 3.01 | Major Histocompatibility Complex, Class I, A | 0.46 |  |

Variants were excluded if there were more than 10% of genotypes missing in the controls and cases or if the heterozygote count was smaller than both homozygote counts in controls and cases. If a subject was not genotyped for a variant then they were assigned the subject-wise average score for that variant. For each subject a gene-wise weighted burden score was derived as the sum of the variant-wise weights, each multiplied by the number of alleles of the variant which the given subject possessed.

Analyses were restricted to the 47 protein-coding genes significant at p < 0.001 in the previous study. For each gene, logistic regression analysis was carried out with hyperlipidaemia as the dependent variable including the first 20 population principal components and sex as covariates and a likelihood ratio test was performed comparing the likelihoods of the models with and without the gene-wise burden score. This is a test for association between the gene-wise burden score and caseness and the statistical significance was summarised as a signed log p value (SLP), which is the log base 10 of the p value given a positive sign if the score is higher in cases and negative if it is higher in controls. Since only 47 genes were analysed, after correcting for multiple testing a gene could be declared statistically significant if it achieved an SLP with absolute value greater than -log10(0.05/47) = 2.97 using the new samples.

Follow-up analyses were performed on all genes individually achieving this significance level and also *ANGPTL4*. For this subset of genes the weighted burden analysis described above was carried out using the whole sample of 106,091 cases and 363,674 controls. Additionally, for each subject a count was obtained of the number of variants they carried falling into particular broad annotation categories, such as LOF, protein altering, etc. The full list of these categories is shown in Supplementary Table 1. These counts were entered into a multiple logistic regression analysis with hyperlipidaemia as the dependent variable and again including sex and 20 principal components as covariates in order to elucidate the contribution of different types of variant to the overall evidence for association. The odds ratios (ORs) associated with each category were estimated along with their standard errors and the Wald statistic was used to obtain a p value. This p value was converted to an SLP, again with the sign being positive if the OR was greater than 1, indicating that variants in that category tended to increase risk.

Data manipulation and statistical analyses were performed using GENEVARASSOC, SCOREASSOC and R (R Core Team, 2014; Curtis, 2016, 2020b).

## Results

Table 1 shows the results of the primary analysis. Three of the four protein-coding genes which reach exome-wide significance in the earlier study show convincing evidence of association with hyperlipidaemia, *LDLR*, *PCSK9* and *ANGPTL3*, with SLPs of 87.28, -29.08 and -7.48 respectively. However the other gene, *IFITM5*, shows no evidence of association, with SLP of only 0.44, so it seems reasonable to conclude that the original results for this gene represented a type 1 error. Of the remaining genes which were originally significant at p < 0.001, most show no evidence of association in this new sample and can be dismissed as chance findings. However *ABCG5*, *APOC3* and *NPC1L1* all produce SLPs which are statistically significant after correcting for testing 47 genes, with values of 5.08, -11.01 and - 3.42. These six genes were carried forward for secondary analyses along with *ANGPTL4* which was considered to be of interest because of its similarity to *ANGPTL3*, even though it achieved SLPs of only -3.66 in the first sample and -1.95 in the second.

The original study considered 22,642 genes, meaning that for a gene-wise result to be considered exome-wide significant the magnitude of the SLP obtained should exceed - log10(0.05/22642) = 5.66. For the seven genes carried forward, the results of weighted burden analysis in the entire sample of 106,091 cases and 363,674 controls are also shown in Table 1 and it can be seen that all six of the genes which produced results which were statistically significant after multiple testing in the second sample also produce results which would be regarded as exome-wide significant in the full sample. However the SLP for *ANGPTL3* in the combined sample is only -4.79.

In order to gain insights into the effects of different categories of variant within these seven genes of interest, counts for variants of each category in each subject were entered into multiple logistic regression analysis along with sex and 20 principal components as covariates. These results are shown in Tables 2 - 4 and are summarised briefly as follows.

Variants in LDLR (SLP = 156.81) and ABCG5 (SLP = 6.95) increase risk of hyperlipidaemia and results for each variant category are shown in Table 2. Table 2A shows the results for *LDLR* and it can be seen that LOF variants are associated with hyperlipidaemia risk with OR > 20. 113 participants carry a LOF variant and all but 16 of these are cases. Of note, there are also 19 subjects who carry an inframe indel and all but 2 of these are also cases, again yielding an OR over 20 though with a wide confidence interval. Detailed inspection of these results reveals that they are driven by two inframe deletions, 19:11105556ATGG>A (rs121908027) which is carried by 8 participants who are all cases and 19:11116925ACGG>A (rs1221971156) which is carried by 7 participants, 6 of whom are cases. The first of these, rs121908027, is reported to the be most common familial hyperlipidaemia (FH) mutation in Ashkenazi Jews and was found in 35% of FH families in Israel (Meiner *et al*., 1991). As well as the large effect of these LOF and indel variants there is statistically significant evidence for an overall small effect on risk of the much commoner variants in the "Protein altering" category (consisting mostly of nonsynonymous variants) with OR of 1.12 and a further modest increase in risk if these are annotated as deleterious by SIFT and/or possibly or probably damaging by PolyPhen, with ORs of 1.44, 1.27 and 1.61. While all these categories of variant are associated with increased risk of hyperlipidaemia, the category "Splice region" is actually associated with reduced risk, with OR of 0.82 and SLP of -19.81. This result is driven by 19:11120527G>A (rs72658867), which has MAF 0.0126 in controls and 0.0096 in cases and which has previously been reported to lower HDL cholesterol and to be protective against coronary artery disease (Gretarsdottir *et al*., 2015).

**Table 2.** Results from logistic regression analysis showing the contribution different categories of variant within each gene make to risk of hyperlipidaemia. Odds ratios for each category are estimated including principal components and sex as covariates.

**Table 2A.** Results for *LDLR*.

| Variant category | Number of separate variants | Total count in controls | Mean count in controls | Total count in cases | Mean count in cases | OR (95% confidence interval) | SLP |
| --- | --- | --- | --- | --- | --- | --- | --- |
| Intronic, etc | 1116 | 62789 | 0.172651 | 18763 | 0.176854 | 1.03 (1.01 - 1.04) | 2.80 |
| 5 prime UTR | 46 | 172 | 0.000473 | 44 | 0.000415 | 0.89 (0.63 - 1.26) | -0.30 |
| Synonymous | 283 | 5947 | 0.016353 | 1639 | 0.015449 | 0.93 (0.88 - 0.98) | -2.02 |
| Splice region | 81 | 15506 | 0.042638 | 4022 | 0.037911 | 0.82 (0.79 - 0.86) | -19.81 |
| 3 prime UTR | 27 | 1248 | 0.003433 | 342 | 0.003225 | 0.95 (0.84 - 1.08) | -0.39 |
| Protein altering | 624 | 11496 | 0.031611 | 4287 | 0.040409 | 1.12 (1.06 - 1.19) | 4.43 |
| Indel, etc | 6 | 2 | 0.000005 | 17 | 0.000160 | 25.66 (5.67 - 116.17) | 8.96 |
| LOF | 44 | 16 | 0.000044 | 97 | 0.000914 | 23.48 (13.63 - 40.46) | 30.39 |
| SIFT deleterious | 329 | 1736 | 0.004774 | 1148 | 0.010821 | 1.44 (1.26 - 1.65) | 7.15 |
| PolyPhen possibly damaging | 123 | 964 | 0.002651 | 472 | 0.004449 | 1.27 (1.10 - 1.46) | 3.16 |
| PolyPhen probably damaging | 197 | 1299 | 0.003572 | 898 | 0.008465 | 1.61 (1.39 - 1.86) | 9.80 |

Table 2B shows the results for *ABCG5* and it can be seen that although a few hundred participants carry LOF variants these do not appear to have any strong effect on hyperlipidaemia risk with an OR of 1.2 which is not statistically significant. Instead, the signal for this gene seems to be driven largely by the "Splice region category", with OR of 1.44 and SLP of 6.20. Although there are 74 variants in this category, the result seems to be mainly driven by three variants which are somewhat commoner in cases, 2:43813316A>C (rs114780578), 2:43822939G>C (rs370895243) and 2:43825025A>T (rs201469377). The category "Protein altering" yields an OR of 1.05 and an SLP of 2.07 but there is no suggestion that nonsynonymous variants recognised as more severe by SIFT or PolyPhen are associated with increased risk. This result may be largely driven by 2:43813208T>C (rs140374206) which had frequency 0.006049 in controls and 0.006246 in cases and which has previously been reported to be associated with raised non-HDL cholesterol and increased risk of gallstones (Helgadottir *et al*., 2020).

Variants in *PCSK9* (SLP = -48.57), *NPC1L1* (SLP = -7.60) and *APOC3* (SLP = -13.19) are protective against hyperlipidaemia and their results detailed are shown in Table 3. As can be seen in Table 3A, LOF variants in *PCSK9* reduce hyperlipidaemia risk, with OR of 0.39. On average, protein altering variants in general have a mild effect on lowering risk, with OR of 0.92, but those which are additionally annotated as deleterious by SIFT have a larger effect, with OR 0.69, whereas there is no additional effect associated with being characterised as possibly or probably damaging by PolyPhen.

**Table 2B.** Results for *ABCG5*.

| Variant category | Number of separate variants | Total count in controls | Mean count in controls | Total count in cases | Mean count in cases | OR (95% confidence interval) | SLP |
| --- | --- | --- | --- | --- | --- | --- | --- |
| Intronic, etc | 1370 | 30573 | 0.084068 | 9093 | 0.085713 | 1.01 (0.99 - 1.03) | 0.49 |
| 5 prime UTR | 77 | 517 | 0.001422 | 158 | 0.001489 | 0.99 (0.83 - 1.19) | -0.03 |
| Synonymous | 222 | 3828 | 0.010526 | 1218 | 0.011481 | 1.04 (0.97 - 1.11) | 0.52 |
| Splice region | 74 | 674 | 0.001854 | 271 | 0.002555 | 1.44 (1.24 - 1.66) | 6.20 |
| 3 prime UTR | 7 | 18 | 0.000049 | 2 | 0.000019 | 0.35 (0.08 - 1.56) | -0.55 |
| Protein altering | 520 | 17153 | 0.047166 | 5476 | 0.051619 | 1.05 (1.01 - 1.09) | 2.07 |
| Indel, etc | 1 | 0 | 0.000000 | 2 | 0.000019 |  | 1.29 |
| LOF | 58 | 368 | 0.001012 | 128 | 0.001207 | 1.20 (0.98 - 1.48) | 1.12 |
| SIFT deleterious | 290 | 5381 | 0.014797 | 1831 | 0.017259 | 1.08 (0.96 - 1.23) | 0.69 |
| PolyPhen possibly damaging | 96 | 2104 | 0.005785 | 694 | 0.006542 | 0.98 (0.85 - 1.13) | -0.10 |
| PolyPhen probably damaging | 161 | 2321 | 0.006382 | 813 | 0.007663 | 1.05 (0.92 - 1.21) | 0.33 |

**Table 3.** Results of variant category analysis for *PCSK9, NPC1L1* and *APOC3*.

**Table 3A.** Results for *PCSK9*.

| Variant category | Number of separate variants | Total count in controls | Mean count in controls | Total count in cases | Mean count in cases | OR (95% confidence interval) | SLP |
| --- | --- | --- | --- | --- | --- | --- | --- |
| Intronic, etc | 695 | 13345 | 0.036694 | 4024 | 0.037926 | 1.03 (1.00 - 1.07) | 1.21 |
| 5 prime UTR | 48 | 657 | 0.001807 | 186 | 0.001753 | 0.98 (0.83 - 1.16) | -0.09 |
| Synonymous | 230 | 16789 | 0.046165 | 4999 | 0.047120 | 0.99 (0.96 - 1.03) | -0.15 |
| Splice region | 55 | 698 | 0.001919 | 223 | 0.002102 | 1.06 (0.91 - 1.24) | 0.36 |
| 3 prime UTR | 40 | 5701 | 0.015676 | 1719 | 0.016206 | 1.02 (0.97 - 1.08) | 0.34 |
| Protein altering | 537 | 7905 | 0.021737 | 1891 | 0.017824 | 0.92 (0.86 - 0.98) | -1.93 |
| Indel, etc | 10 | 2620 | 0.007205 | 758 | 0.007145 | 1.00 (0.92 - 1.08) | -0.03 |
| LOF | 72 | 862 | 0.002370 | 94 | 0.000886 | 0.39 (0.31 - 0.48) | -17.41 |
| SIFT deleterious | 244 | 3354 | 0.009223 | 655 | 0.006174 | 0.69 (0.59 - 0.81) | -5.46 |
| PolyPhen possibly damaging | 70 | 616 | 0.001694 | 177 | 0.001668 | 1.18 (0.98 - 1.43) | 1.12 |
| PolyPhen probably damaging | 157 | 2408 | 0.006621 | 475 | 0.004477 | 1.01 (0.85 - 1.21) | 0.05 |

The results in Table 3B show that the overall signal for *NPC1L1* is mainly due to LOF variants, with OR 0.64 and SLP of -5.20, with possibly some additional contribution from variants annotated as deleterious by SIFT, which have OR 0.89 and SLP -1.76. A similar scenario is seen for *APOC3* in Table 3C, with LOF variants have OR of 0.68 and SLP of - 11.22 but with other categories not showing clear evidence of association.

**Table 3B.** Results for *NPC1L1*.

| Variant category | Number of separate variants | Total count in controls | Mean count in controls | Total count in cases | Mean count in cases | OR (95% confidence interval) | SLP |
| --- | --- | --- | --- | --- | --- | --- | --- |
| Intronic, etc | 1197 | 30977 | 0.085178 | 9035 | 0.085162 | 1.00 (0.97 - 1.02) | -0.13 |
| 5 prime UTR | 24 | 853 | 0.002346 | 275 | 0.002592 | 1.14 (0.99 - 1.31) | 1.19 |
| Synonymous | 442 | 11147 | 0.030651 | 3218 | 0.030333 | 0.98 (0.94 - 1.02) | -0.54 |
| Splice region | 74 | 605 | 0.001664 | 164 | 0.001546 | 0.94 (0.79 - 1.13) | -0.30 |
| 3 prime UTR | 31 | 954 | 0.002624 | 282 | 0.002659 | 0.96 (0.84 - 1.10) | -0.25 |
| Protein altering | 926 | 28236 | 0.077642 | 7764 | 0.073184 | 0.98 (0.96 - 1.00) | -0.94 |
| Indel, etc | 13 | 43 | 0.000118 | 16 | 0.000151 | 1.31 (0.72 - 2.38) | 0.44 |
| LOF | 100 | 661 | 0.001818 | 123 | 0.001160 | 0.64 (0.53 - 0.78) | -5.20 |
| SIFT deleterious | 507 | 8949 | 0.024607 | 2366 | 0.022302 | 0.89 (0.80 - 0.98) | -1.76 |
| PolyPhen possibly damaging | 143 | 5970 | 0.016416 | 1532 | 0.014441 | 1.00 (0.90 - 1.11) | -0.01 |
| PolyPhen probably damaging | 325 | 2798 | 0.007694 | 817 | 0.007701 | 1.15 (1.01 - 1.30) | 1.52 |

**Table 3C.** Results for *APOC3*.

| Variant category | Number of separate variants | Total count in controls | Mean count in controls | Total count in cases | Mean count in cases | OR (95% confidence interval) | SLP |
| --- | --- | --- | --- | --- | --- | --- | --- |
| Intronic, etc | 134 | 10194 | 0.028031 | 2817 | 0.026555 | 0.95 (0.91 - 1.01) | -1.13 |
| 5 prime UTR | 2 | 1 | 0.000003 | 1 | 0.000009 | 3.51 (0.20 - 62.29) | 0.4 |
| Synonymous | 31 | 207 | 0.000569 | 48 | 0.000452 | 0.81 (0.59 - 1.12) | -0.7 |
| Splice region | 11 | 119 | 0.000327 | 37 | 0.000349 | 1.06 (0.72 - 1.55) | 0.11 |
| 3 prime UTR | 33 | 199 | 0.000547 | 56 | 0.000528 | 1.01 (0.75 - 1.37) | 0.03 |
| Protein altering | 69 | 568 | 0.001562 | 136 | 0.001282 | 1.06 (0.77 - 1.45) | 0.13 |
| Indel, etc | 2 | 48 | 0.000132 | 8 | 0.000075 | 0.58 (0.27 - 1.25) | -0.81 |
| LOF | 10 | 1993 | 0.005480 | 402 | 0.003789 | 0.68 (0.61 - 0.76) | -11.22 |
| SIFT deleterious | 30 | 416 | 0.001144 | 89 | 0.000839 | 0.47 (0.22 - 1.02) | -1.29 |
| PolyPhen possibly damaging | 14 | 40 | 0.000110 | 18 | 0.000170 | 2.77 (1.15 - 6.68) | 1.69 |
| PolyPhen probably damaging | 10 | 325 | 0.000894 | 66 | 0.000622 | 1.35 (0.63 - 2.92) | 0.37 |

Variants in *ANGPTL3* (SLP = -12.68) and *ANGPTL4* (SLP = -4.79) also appear to be protective against hyperlipidaemia, although the result for *ANGPTL4* is not exome-wide significant. Nevertheless, as the products of both genes modulate the activity of lipoprotein lipase and as inactivating variants in both genes have previously been shown to be associated with hypolipidaemia, it seems appropriate to present the detailed results for both, as shown in Table 4 (Dron and Hegele, 2016; Wang and Musunuru, 2019). For *ANGPTL3* the signal is again mainly due to LOF variants, with OR of 0.59 and SLP of -8.36, but it can be seen that splice region variants also have OR of 0.69 and SLP of -4.70. This latter result is driven by 1:62598067T>C (rs372257803) which has MAF 0.00100 in controls and 0.00069 in cases and which has been previously reported to be associated with lower non-HLD cholesterol and triglycerides (Helgadottir *et al*., 2016). The results for *ANGPTL4* are not statistically significant after correction for multiple testing but it can be seen that they are consistent with the possibility that LOF variants lower hyperlipidaemia risk modestly, with OR 0.78 and SLP -1.84.

**Table 4.** Results of variant category analysis for *ANGPTL3* and *ANGPTL4*.

**Table 4A.** Results for *ANGPTL3*.

| Variant category | Number of separate variants | Total count in controls | Mean count in controls | Total count in cases | Mean count in cases | OR (95% confidence interval) | SLP |
| --- | --- | --- | --- | --- | --- | --- | --- |
| Intronic, etc | 347 | 8024 | 0.022064 | 2585 | 0.024370 | 1.02 (0.96 - 1.08) | 0.29 |
| 5 prime UTR | 25 | 3604 | 0.009909 | 1104 | 0.010408 | 0.99 (0.87 - 1.14) | -0.04 |
| Synonymous | 103 | 4397 | 0.012091 | 1322 | 0.012461 | 0.96 (0.86 - 1.08) | -0.28 |
| Splice region | 22 | 803 | 0.002208 | 161 | 0.001518 | 0.69 (0.58 - 0.82) | -4.70 |
| 3 prime UTR | 34 | 2019 | 0.005550 | 560 | 0.005280 | 0.97 (0.88 - 1.06) | -0.32 |
| Protein altering | 265 | 10476 | 0.028806 | 2945 | 0.027759 | 0.94 (0.85 - 1.03) | -0.77 |
| Indel, etc | 6 | 93 | 0.000256 | 20 | 0.000189 | 0.69 (0.42 - 1.15) | -0.84 |
| LOF | 56 | 835 | 0.002296 | 143 | 0.001348 | 0.59 (0.49 - 0.70) | -8.36 |
| SIFT deleterious | 137 | 6886 | 0.018935 | 1930 | 0.018192 | 0.97 (0.86 - 1.08) | -0.26 |
| PolyPhen possibly damaging | 41 | 7682 | 0.021123 | 2175 | 0.020501 | 1.07 (0.94 - 1.23) | 0.54 |
| PolyPhen probably damaging | 69 | 708 | 0.001947 | 193 | 0.001819 | 1.00 (0.81 - 1.24) | 0.01 |

**Table 4B.**
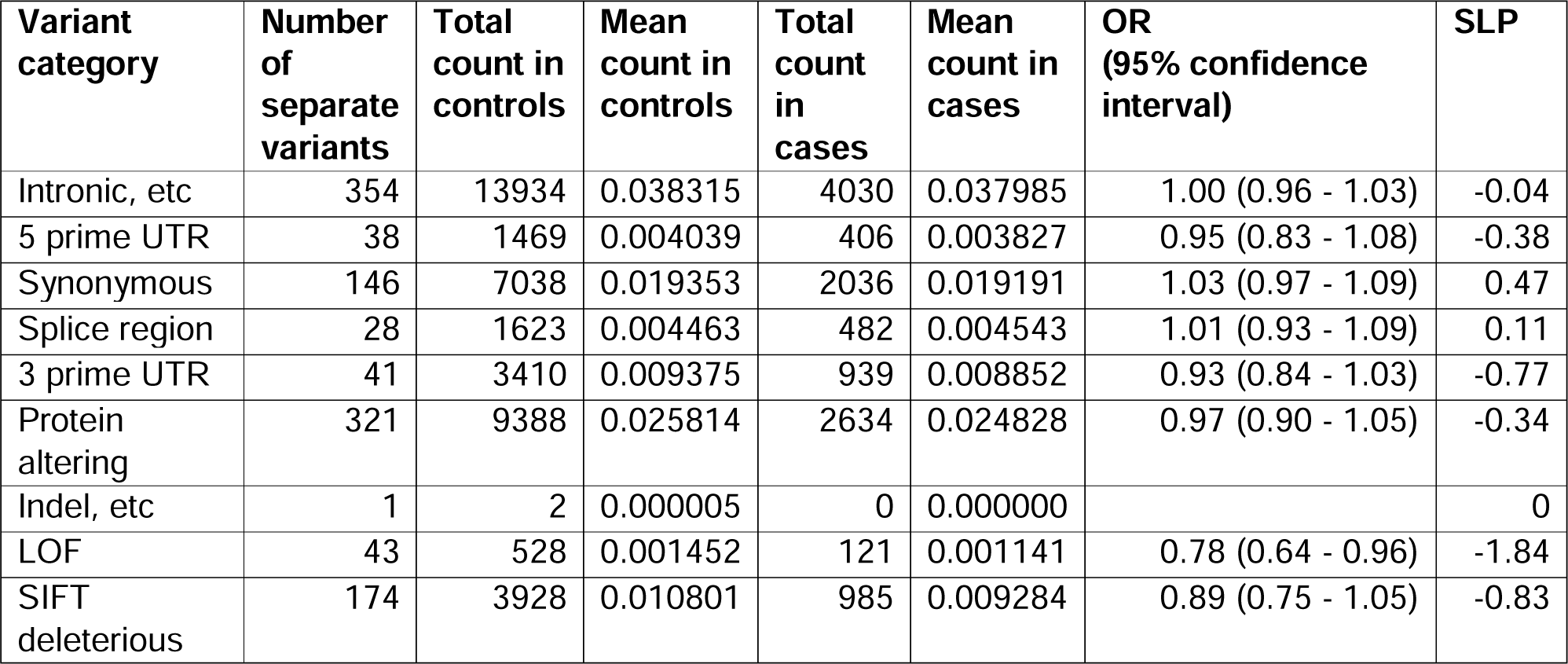
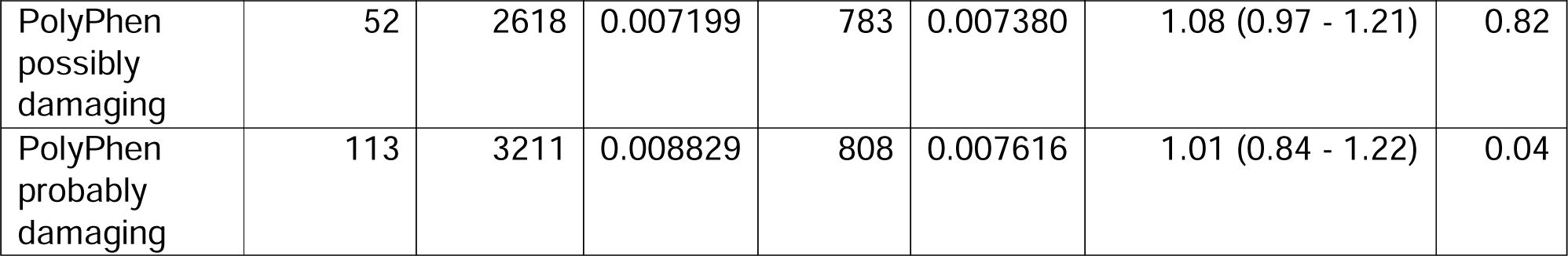
Results for *ANGPTL4*.

## Discussion

As mentioned above, this dataset has been used for previous two studies testing for association between exome sequence variance with a very large number of phenotypes, which for convenience we can refer to as the Regeneron and AstraZeneca studies (Backman *et al*., 2021; Wang *et al*., 2021). The Regeneron study carried out a variety of single variant and gene-wise burden tests on 3,994 health-related traits to produce a total of about 2.3 billion tests, yielding a critical p value of 2.18LJ×LJ10^−11^ (corresponding to SLP = 10.66)) and reported 8,865 significant associations which are presented in their Supplementary Data 2 (Backman *et al*., 2021). This reports significant gene-wise associations of measured cholesterol and/or LDL levels for all seven of the genes implicated in the present study. However the only genes reported to be associated with clinical hyperlipidaemia, as indicated by taking lipid lowering medication or having a hyperlipidaemia diagnosis recorded, were *LDLR*, *PCSK9* and *APOC3*. For the AstraZeneca study, all gene-wise and variant-wise associations with 17,361 binary and 1,419 quantitative phenotypes are reported on the AstraZeneca PheWAS Portal at https://azphewas.com/ (Wang *et al*., 2021). This was accessed to find the most significant p value for any analysis of each of these genes with the phenotype "Union#E78#E78 Disorders of lipoprotein metabolism and other lipidaemias" and Table 5 shows the results obtained compared with those for the current study. This shows that, for every gene, the AstraZeneca results provide less support for association than the present study and in particular both *ABCG5* and *NPC1L1* would be regarded as exome-wide significant in the present study. While, these two genes were both previously shown to influence lipid levels, the current study implicates variants in these genes as impacting the risk of developing clinically relevant hyperlipidaemia, suggesting that such variants could be included when drawing up genetically informed individual risk profiles.

**Table 5.** Comparison of results from current study to those reported for the AstraZeneca study. The results for the AstraZeneca study are displayed as the equivalent SLP for the most significant result reported for that gene with the phenotype "Union#E78#E78 Disorders of lipoprotein metabolism and other lipidaemias".

| Gene | SLP for combined sample in current study | SLP for AstraZeneca study |
| --- | --- | --- |
| *LDLR* | 156.81 | 70.73 |
| *ABCG5* | 6.95 | 3.94 |
| *PCSK9* | -43.57 | -17.12 |
| *NPC1L1* | -7.60 | 5.21 |
| *APOC3* | -13.19 | -12.47 |
| *ANGPTL3* | -12.68 | -11.66 |
| *ANGPTL4* | -4.79 | 2.08 |

Of the four genes in which variants impairing function are associated with reduced risk of hyperlipidaemia, two are already established drug targets. The product of *NPC1L1*, which is essential for intestinal sterol absorption, is the molecular target of ezetimibe, a cholesterol absorption inhibitor that lowers blood cholesterol (Betters and Yu, 2010). The product of *ANGPTL3* is the target of evinacumab, a human monoclonal antibody designed to treat hypercholesterolaemia (Wang and Musunuru, 2019; Doggrell, 2020). The well-established evidence for *PCSK9* and *APOC3* variants as being protective is fuelling research into developing strategies to find novel ways to antagonise PCSK9 and lower apoC-III (Taskinen, Packard and Borén, 2019; Kuzmich *et al*., 2022).

The detailed analyses of variant categories provide insights into the magnitude of effects of different kinds of variant in different genes, along with information about their cumulative frequencies. One feature of note is the heterogeneity of effects between genes. For example, for *LDLR* nonsynonymous variants classified as probably damaging by PolyPhen are more strongly implicated and have a larger effect size than variants classified as deleterious by SIFT, but for *PCSK9* this is not the case and only SIFT deleterious variants have an effect. For *ANGPTL3* neither SIFT nor PolyPhen is helpful for identifying risk-associated variants. For most genes LOF variants have the largest effect size but this is not the case for *ABCG5* in which LOF variants do not clearly have any effect and the signal is instead driven by specific splice region variants. This inconsistency of effects of different annotations across genes was noted in a previous study and seems to imply that there is no universal system for weighting variants which would perform optimally for all genes (Curtis, 2022).

The present study quantifies the effect on hyperlipidaemia risk of naturally occurring variation within these genes, along with their cumulative frequencies in a sample broadly representative of the population. The fact that people with severe, early onset cardiovascular disease might have been less likely to survive to be recruited into UK Biobank may mean that the magnitude of effect of some variants on risk is somewhat underestimated, but this does not seem likely to be a major consideration. Variants with large effect sizes are very rare. Around 10,000 participants carry a variant in a category with a moderate effect on risk, i.e. with OR below 0.7 or above 1.4, but it is debatable how relevant such effects would be for quantifying individual risk in the context of personalised medicine. It seems that the main value of identifying coding variants associated with risk is to clarify the pathophysiological mechanisms influencing risk of hyperlipidaemia in order to support the development of novel therapeutic approaches.

### Conflicts of interest

The author declares he has no conflict of interest.

### Data availability

The raw data is available on application to UK Biobank. Detailed results with variant counts cannot be made available because they might be used for subject identification. Relevant derived variables including principal components and variant annotations will be deposited in UK Biobank. Scripts and software used to carry out the analyses are available at https://github.com/davenomiddlenamecurtis.

## Acknowledgments

This research has been conducted using the UK Biobank Resource. The author wishes to acknowledge the staff supporting the High Performance Computing Cluster, Computer Science Department, University College London.

## Funding statement

No external funding was received for this work.

---

**Supplementary Table 1.** The table shows the broad categories used for variant category specific analyses along with the annotations produced by VEP which were grouped into each category.

| Category | VEP annotation |
| --- | --- |
| Intronic etc. | feature_truncation, regulatory_region_variant, feature_elongation, regulatory_region_amplification, regulatory_region_ablation, TF_binding_site_variant, TFBS_amplification, TFBS_ablation, downstream_gene_variant, upstream_gene_variant, non_coding_transcript_variant, NMD_transcript_variant, intron_variant, non_coding_transcript_exon_variant |
| Five prime UTR | 5_prime_UTR_variant |
| Synonymous | synonymous_variant |
| Splice region | splice_region_variant |
| Three prime UTR | 3_prime_UTR_variant |
| Protein altering | protein_altering_variant, missense_variant |
| Indel etc. | inframe_deletion, inframe_insertion, transcript_amplification |
| LOF | frameshift_variant, stop_gained, transcript_ablation, splice_donor_variant, splice_acceptor_variant |
| SIFT deleterious | deleterious |
| PolyPhen possibly damaging | possibly_damaging |
| PolyPhen probably damaging | probably_damaging |

## Notes

### Competing Interest Statement

The authors have declared no competing interest.

### Funding Statement

This study did not receive any funding

### Author Declarations

UK Biobank had obtained ethics approval from the North West Multi-centre Research Ethics Committee which covers the UK (approval number: 11/NW/0382) and had obtained informed consent from all participants. The UK Biobank approved an application for use of the data (ID 51119) and ethics approval for the analyses was obtained from the UCL Research Ethics Committee (11527/001).

